# Bioinformatic approaches to draft the viral genome sequence of Canary Islands cases related to the multi-country Monkeypox virus 2022-outbreak

**DOI:** 10.1101/2022.11.08.22282035

**Authors:** Adrián Muñoz-Barrera, Laura Ciuffreda, Julia Alcoba-Florez, Luis A. Rubio-Rodríguez, Héctor Rodríguez-Pérez, Helena Gil-Campesino, Diego García-Martínez de Artola, Josmar Salas-Hernández, Julia Rodríguez-Núñez, Antonio Íñigo-Campos, Oscar Díez-Gil, Rafaela González-Montelongo, Agustín Valenzuela-Fernández, José M. Lorenzo-Salazar, Carlos Flores

## Abstract

During May 2022, the World Health Organization declared community transmission of monkeypox (MPXV) due to a multi-country outbreak. In Europe, several cases of this outbreak were detected in the Canary Islands (Spain). Here we describe the combination of DNA sequencing and bioinformatic approaches, including methods for *de novo* genome assembly and short- and long-read technologies, used to reconstruct the first MPXV genome isolated in the Canary Islands on the 31^st^ of May 2022 from a male adult patient with mild symptoms. We obtained the best results using a reference-based approach with short reads, evidencing 46-67 nucleotide variants against sequences from the 2018-2019 outbreak, and placing the sequence in the B.1 clade. This study demonstrates the potential of metagenomics sequencing for rapid and precise pathogen identification.

## Introduction

Monkeypox virus (MPXV) is a zoonotic Orthopoxvirus (OPV) (family *Poxviridae*) [1,2], endemic to West and Central Africa [3,4]. MPXV has been described in humans in Central and Western Africa (occurring mainly in tropical forest areas of Central Africa) as well as in other parts of the world [5–10]. Around the 13^th^ of May 2022, MPXV cases were reported in several countries and WHO declared community transmission of the virus [11]. Most reported cases so far have been presented through sexual health or other health services and have involved mainly men who have sex with men [12,13]. By early June 2022, 129 viral genomes had been deposited at GISAID [14] with 46 SNPs shared by all these sequences and differing from the viral genome sequences from the 2018-2019 MPXV outbreak [15]. Preliminary data from polymerase chain reaction (PCR) assays indicate that these MPXV strains detected in Europe and other non-endemic areas belong to the West African clade [16].

In Europe, several cases of MPXV infection have been associated with 2022 outbreaks in the Canary Islands and Spain [16]. A few viral sequences from samples collected in Spain have been reported [17]. As of June 2022, a total of 15 positive cases were confirmed in the Canary Islands [18], which have increased to a total of 176 by early November 2022. Here we describe the combination of methodological approaches to obtain the draft sequences of the first MPVX viral genome isolated in the Canary Islands on the 31^st^ of May 2022 from a male adult patient with one week-onset mild symptoms (fever, odynophagia) and presenting at the Emergency Room but not necessitating hospital admission. An expedited description of the case and the resulting draft sequences was publicly posted in mid-June 2022 [19].

## Materials and Methods

### DNA extraction and PCR testing

Viral DNA was extracted at the Hospital Universitario Ntra. Sra. de Candelaria (Santa Cruz de Tenerife, Spain) from five samples (nasopharyngeal swab, lesion crust, and vesicles) from the same patient using the eMAG system (Biomerieux) following manufacturer’s instructions. Virus inactivation was conducted under a biosafety class II cabinet (TELSTAR bio-II-A), following ECDC procedures [20]. Diagnosis of MPXV infection was confirmed using the LightMix Modular Orthopox (Roche) and a real-time PCR assay described elsewhere [21]. This assay yielded threshold cycle values in the range of 17 to 33 in these samples.

### Short- and long-read DNA sequencing

Five independent DNA dual index libraries (one for each sample) were processed at Instituto Tecnológico y de Energías Renovables (ITER) with Nextera XT DNA Library Preparation Kit (Illumina Inc.), following the manufacturer’s recommendations with manual library normalization, and pooled prior to sequencing. The quality of the libraries was assessed with a D1000 ScreenTape kit on the 4200 TapeStation System (Agilent). Library concentrations ranged from 7.4 to 10.4 nM, and showed a fragmentation profile ranging from 721 to 808 bp. The mean fragment size for the sequencing pool was 677 bp as measured with a D1000 High Sensitivity ScreenTape kit (Agilent). Paired-end sequences were obtained on a MiSeq Sequencing System (Illumina Inc.), using the reagent kit v3 chemistry with 150 cycles and an expected throughput of 3.3-3.8 Gb. The pool concentration was 15 pM, and 5% of PhiX Control V3 was used as the internal control.

DNA libraries for nanopore sequencing were also prepared from the sample with the highest yield (taken from a skin lesion exudate) using the Rapid Barcoding kit (SQK-RBK004) from Oxford Nanopore Technologies (ONT). To increase the quantity of the starting material, the protocol used 30 to 45 ng of the DNA extract in 7.5 μl for 12 independently barcoded libraries that were pooled in order to obtain the maximum yield from the run. The pooled libraries were loaded onto an R9.4.1 flow cell and were run in a MinION (ONT) for 42 hours. Basecalling of raw ONT signal data as well as demultiplexing and adapter trimming was carried out using Guppy v.6.0.7 with default parameters and the high-accuracy basecalling model.

### Bioinformatic analyses and assembly comparisons

As the first step, the individual demultiplexed FASTQ pair of Illumina files were interleaved with BBMap (Reformat tool) and then merged into a single interleaved FASTQ file. Then, two different bioinformatic tools were tested to identify and remove the human reads: Kraken 2 [22] and NCBI SRA Human Scrubber v.1.0.2021_05_05 (only used for Illumina sequencing data). The remaining Illumina and ONT reads were subjected to different bioinformatic procedures to obtain draft sequences (**Figure 1**).

**Figure 1.**
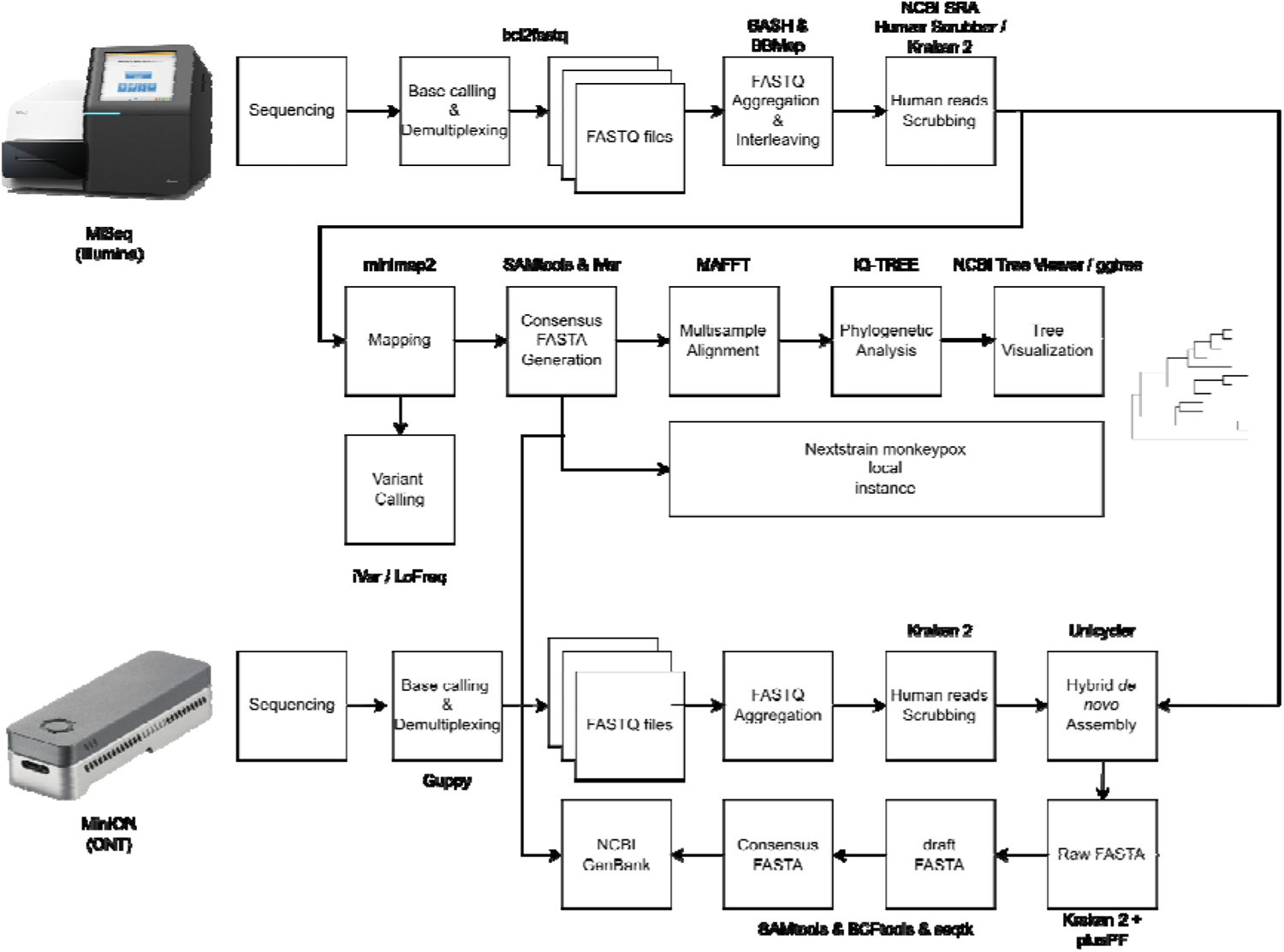
Full bioinformatic pipeline to obtain the MPXV sequences from Illumina-read only and the hybrid *de novo* assembly to infer phylogenetic relationships with other MPXV viral genomes available from public repositories.

On the one hand, a reference-based analysis was conducted with Illumina unclassified reads that were mapped to the MPVX viral genome MPXV-UK_P2 (GenBank MT903344.1) by means of three alternative aligners: minimap2 v.2.24-r1122 [23], BWA-MEM v.0.7.17 [24], and Bowtie2 v.2.4.5 [25]. At this stage, duplicate metrics from PICARD v.2.18.7 [26], and coverage metrics from SAMtools v.1.6 [27] and mosdepth v.0.3.3 [28] were obtained from the remaining interleaved paired-end reads. Variant calling was carried out with two alternative algorithms: iVar v1.3.1 [29] and LoFreq v.2.1.5 [30] using default parameters against the MPXV MT903344.1 genome. For downstream analyses, a consensus sequence was obtained by piping a SAMtools v.1.6 pileup with iVar v.1.3.1 consensus as described elsewhere [31].

On the other hand, a hybrid *de novo* assembly was obtained by combining the filtered Illumina and ONT reads using custom script based on the Unicycler v.0.5.0 [32] assembler. Bandage v.0.9.0 [33] was used to visualize the resulting contigs in the assembly. A refined version of this hybrid *de novo* assembly was obtained after running Kraken 2 v.2.1.2 with PlusPF database to remove non-viral assembled contigs. The consensus sequence of this assembly was obtained by mapping resulting contigs to MPXV MT903344.1 genome and piping SAMtools v.1.6 pileup with BCFtools v.1.6 and seqtk v.1.3-r106.

Finally, the two selected consensus sequences (Illumina-only and hybrid *de novo* assembly) were compared against the MT903344.1 as the reference genome with QUAST v.5.0.2 [34].

### Phylogenetic analysis

The most complete consensus sequence resulting from the previous stage was aligned with 126 MPXV sequences downloaded from NCBI GenBank (**Table S1**) using MAFFT v.7.505 [35]. A phylogenetic analysis was performed using both IQ-TREE v.2.2.0.3 [36] with the K3Pu+F+I model as the best-predicted model and default parameters, and using a local instance of Nextstrain monkeypox [37].

## Results

The Illumina sequencing run produced 3.88 Gb and 25.5 M reads in total. A mapping of 101,814 and 100,897 Illumina reads was obtained using minimap2 on the NCBI SRA Human Scrubber (mean depth: 38.3x) and Kraken 2 (mean depth: 38.1x), respectively, thus providing equivalent results (Table 1). We estimated as few as 2.81% of duplicated reads and that 99% of the MPXV genome was covered ≥1x, with a fraction of 85% of the viral genome covered at ≥10x. The combination of mapper and variant caller that yielded the smallest and the largest number of nucleotide variants against the reference was BWA+LoFreq (46 nucleotide variants) and minimap2+iVar (67 nucleotide variants), respectively (Table 1). In order to maintain the maximum sequence variability for downstream analyses, the consensus sequence for Illumina-only reads was obtained for the minimap2+iVar combination (total size of 197,221 bp), providing a near-fully complete viral genome (99.91%) against the reference.

**Table 1.**
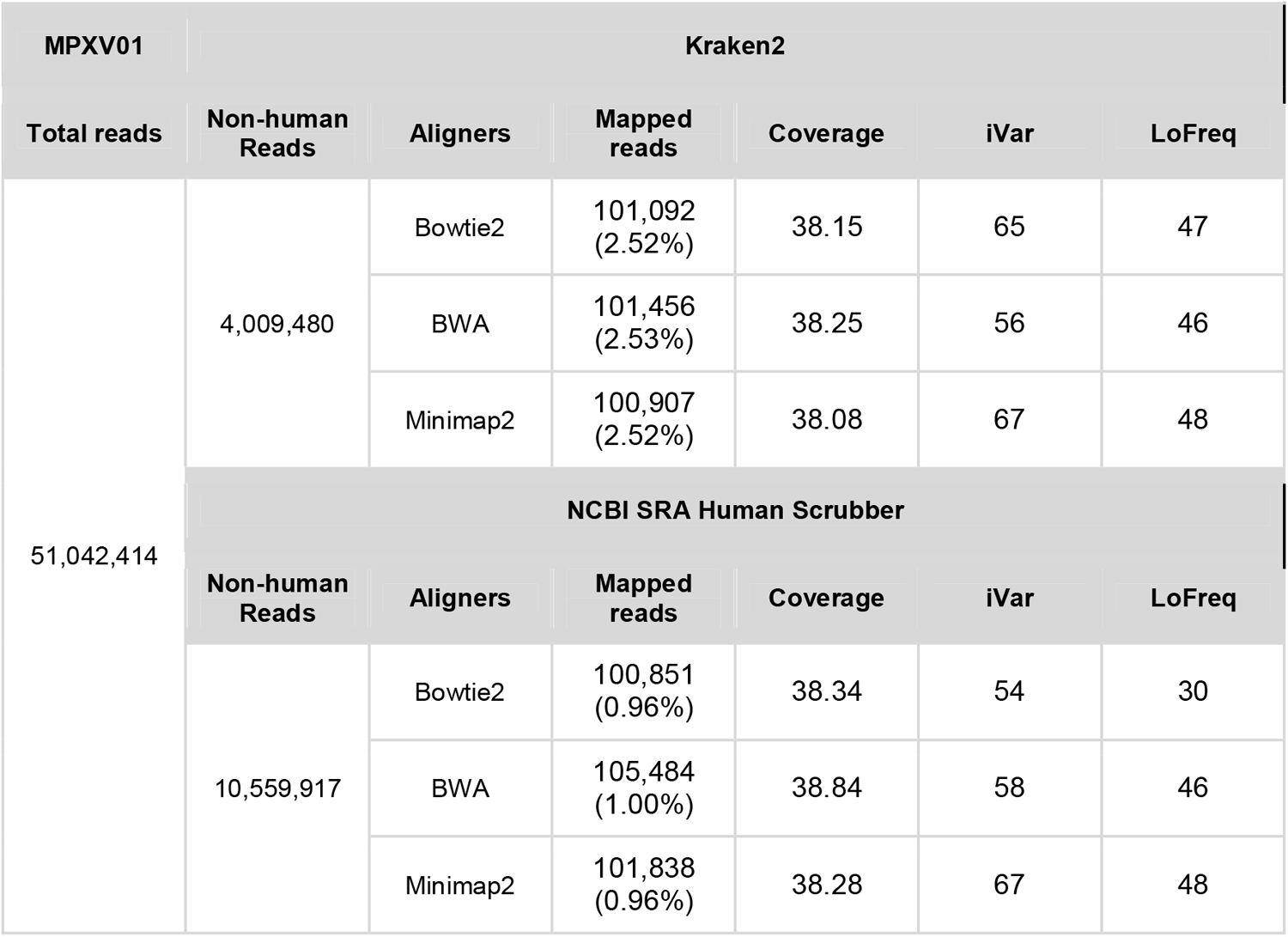
Comparative of mapped Illumina reads and coverage using different aligners (Bowtie2, BWA, Minimap2) and called variants using iVar and LoFreq callers.

The ONT run provided 1.98 Gb and a total of 1.38 M reads, ranging from 499 to 101,895 bp in length, with a mean length of 1,432 bases. ONT sequencing provided 2,246 non-human mapping reads after filtering with Kraken 2, thus, a theoretical viral genome depth of 14.9x. A hybrid *de novo* assembly based on Illumina and ONT Kraken 2-filtered reads was performed and resulted in four contigs (**Figure 2**). Contigs 1 and 2 accounted for 186,315 bp and 4,703 bp (191,018 bp total), respectively, and mapped to Monkeypox virus Zaire-96-I-16. Contigs 3 and 4 spanned 10,530 bp in total but did not map to the MPXV reference. Thus, a consensus sequence was built from this hybrid *de novo* assembly (including only contigs 1 and 2) and the MT903344.1 MPXV reference genome, spanning a total size of 197,222 bp. Note, however, that given that this sequence includes 6,471 undetermined bases, the consensus sequence only covered 96.75% of the reference genome (**Table 2**).

**Table 2.** Assessment of the two MPXV genome assemblies against the MPXV reference.

| Metrics | Hybrid *de novo* assembly<br>(ON782054) | Illumina-only<br>(ON782055) |
| --- | --- | --- |
| # contigs | 1 | 1 |
| Largest contig (bp) | 197,222 | 197,221 |
| Total length (bp) | 197,222 | 197,221 |
| Reference length (bp) | 197,233 | 197,233 |
| GC (%) | 32.93 | 33.02 |
| Reference GC (%) | 33.02 | 33.02 |
| N50 | 197,222 | 197,221 |
| L50 | 1 | 1 |
| # misassemblies | 0 | 0 |
| Genome fraction (%) | 96.75 | 100.00 |
| Duplication ratio | 1.034 | 1.000 |
| # N's per 100 kbp | 3,281.07 | 92.79 |
| # mismatches per 100 kbp | 22.01 | 25.86 |
| # indels per 100 kbp | 0.00 | 1.52 |
| Largest alignment (bp) | 190,825 | 197,221 |
| Total aligned length (bp) | 190,825 | 197,221 |
| NA50 | 190,825 | 197,221 |
| LA50 | 1 | 1 |

**Figure 2.**
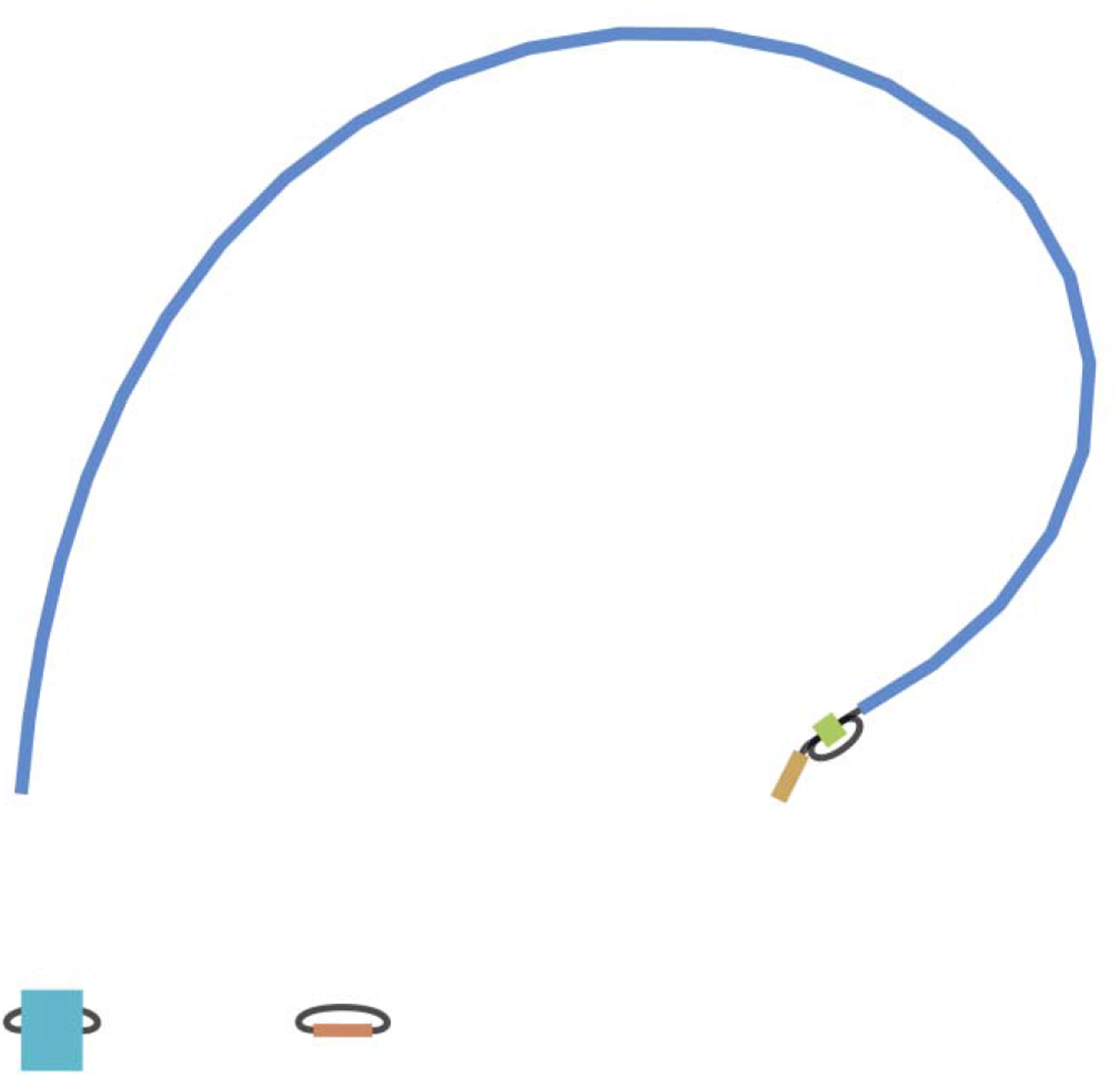
Bandage representation of the hybrid *de novo* assembly based on long-read sequencing technology.

Besides the number of undetermined bases, which were much more in the hybrid *de novo* assembly sequence than in the Illumina-only consensus sequence, we observed small differences overall between the two MPXV genome assemblies obtained (**Table 2**). In brief, the hybrid *de novo* assembly was able to retrieve a slightly lower proportion of the reference genome, although showing a similar GC content and having fewer mismatches than the Illumina-only assembly.

Based on the above findings supporting more completeness for the Illumina-only assembly, we opted for placing this consensus sequence in phylogenetic context (**Figure 3**). This analysis indicated that the draft MPXV genome sequence belongs to the so-called West African clade or B.1 [38,39]. In addition, the closest sequences were related to the Slovenian-MPXV GenBank-released genomes, contributing further evidence of community spread in the present worldwide MPXV outbreak.

**Figure 3.**
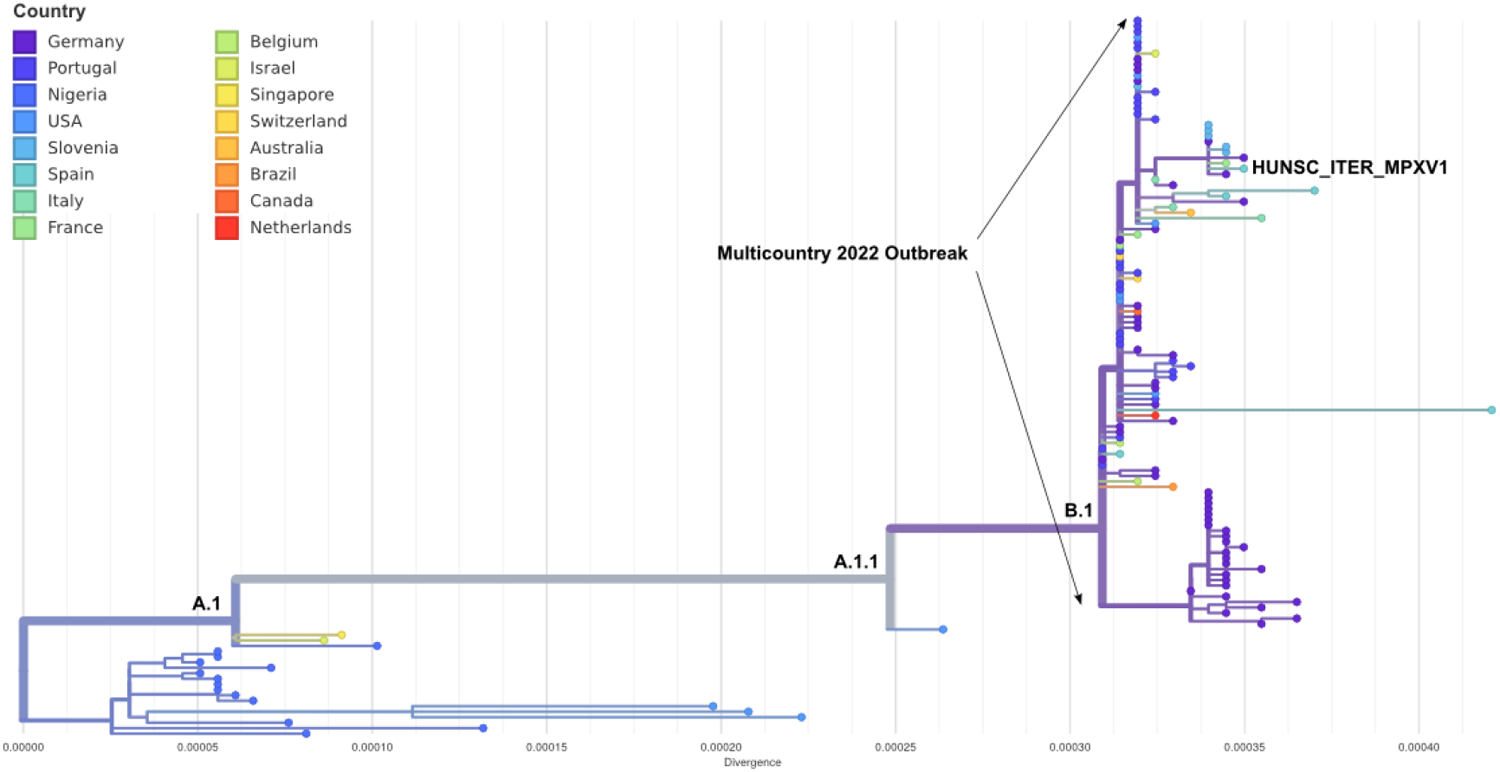
A phylogenetic tree depicting the draft MPXV sequence isolated on May 31, 2022 from a patient from the Canary Islands along with publicly available sequences at the NCBI GenBank.

## Discussion

Here we provide the draft sequences of the first MPVX viral genome isolated in the Canary Islands on 31 May 2022, corresponding to the B.1 clade that was observed across Europe and other non-endemic areas during the outbreak. For that, we have used diverse sequencing and bioinformatic approaches, including methods for *de novo* genome assembly and combining short and long sequencing reads. The best results (higher sequence similarity and higher genome coverage compared to the reference) were obtained using a reference-based approach with Illumina-only reads. With this approximation, and using combinations of mappers and variant callers, between 46 and 67 nucleotide variants were observed when compared to the reference viral genome, which is compatible with the divergence of this new MPXV outbreak from that of 2018-2019 [15].

For this first characterization, we have relied on a metagenomic sequencing approach, providing a low proportion of sequence reads corresponding to the viral genome. In the study, only 0.39% and 0.16% of the reads obtained from the Illumina and ONT runs, respectively, mapped to the MPXV reference. This approach is straightforward and is not as dependent on the viral sequence rearrangements in the outbreaks, as is common for OPVs, demonstrating its value for detecting RNA and DNA viral pathogens in a few hours [40]. However, the costs per sample as well as the viral DNA concentration requirements in the patient samples hinders an extended use. After this work was completed [19], a more sensitive approach based on tiling amplicon sequencing both for Illumina and ONT workflows was developed and validated across a number of laboratories for MPXV [41]. Despite this method being more cost-efficient, a periodic long-read metagenomics sequencing was recommended by the authors to monitor the emergence of viral variants with genomic rearrangements.

Overall, our results provide a proof of concept of the potential of introducing sequencing technologies, and metagenomics in particular, for rapid and precise pathogen diagnosis and surveillance.

## Data Availability

The code used for this study is available at https://github.com/genomicsITER/monkeypox. The Illumina-only and the hybrid de novo assembly consensus MPXV sequences have been released in the NCBI GenBank with accessions ON782054 and ON782055, respectively.

## Ethics statement

The study was conducted at the University Hospital Nuestra Señora de Candelaria (Santa Cruz de Tenerife, Spain) during May 2022. The institutional review board approved the study (ethics approval number: CHUNSC_2022_83).

## Data availability statement

The code used for this study is available at https://github.com/genomicsITER/monkeypox. The Illumina-only and the hybrid *de novo* assembly consensus MPXV sequences have been released in the NCBI GenBank with accessions ON782054 and ON782055, respectively.

## Author contributions

JAF, JMLS and CF designed the study. AMB, LC, JAF, LRR, HRP, RGM, JSH, JRN, AIC, DGM, HGC, ODG, and CF participated in data acquisition. AMB, LRR, HRP, LC, JAF, JMLS and CF performed the analyses and data interpretation. LC, AVF, JMLS and CF wrote the draft of the manuscript. CF obtained funding. All authors contributed in the critical revision and final approval of the manuscript.

## Acknowledgements

We would like to deeply thank the patient participating in the study.

## Supplementary Material

**Table S1.**
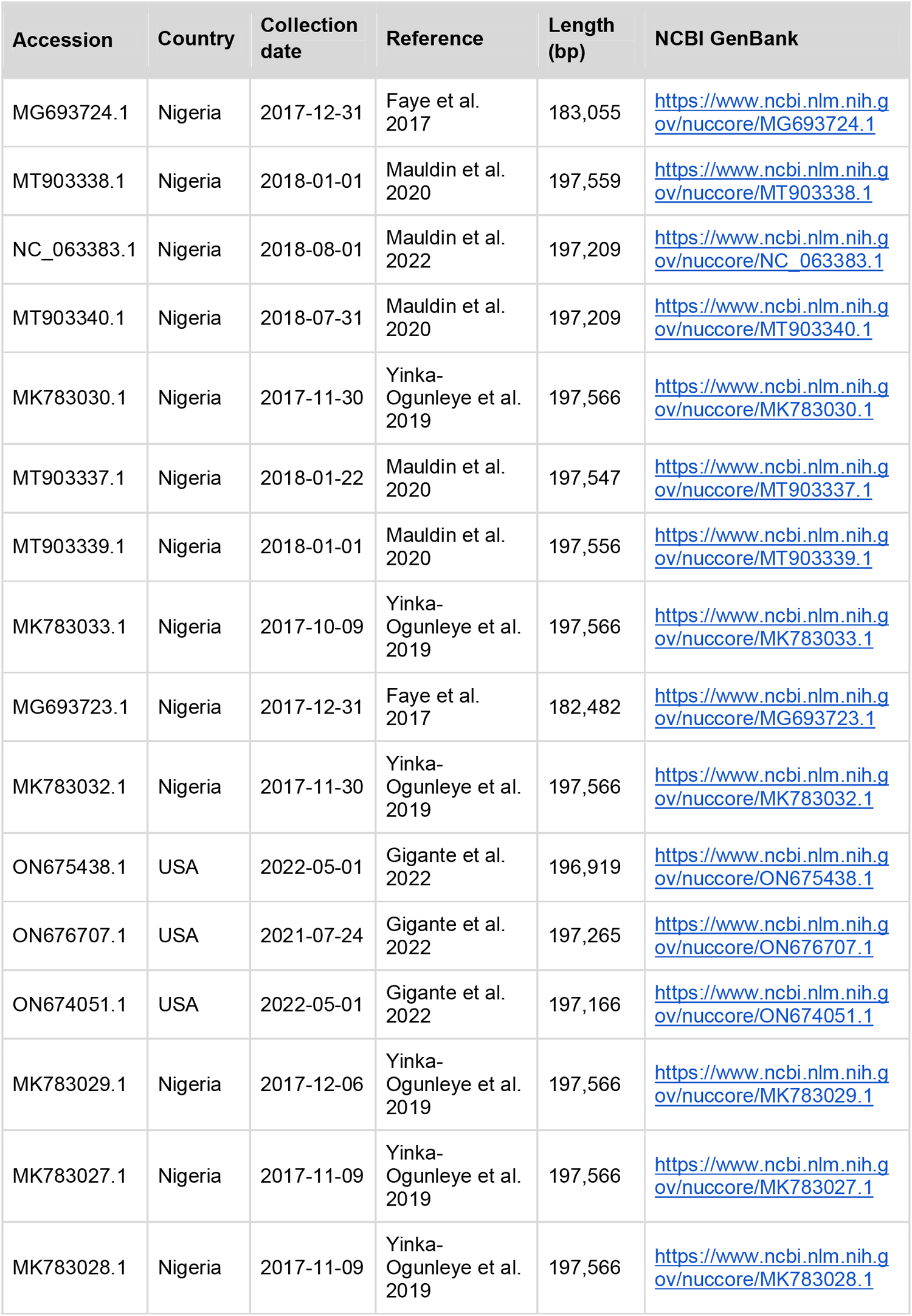
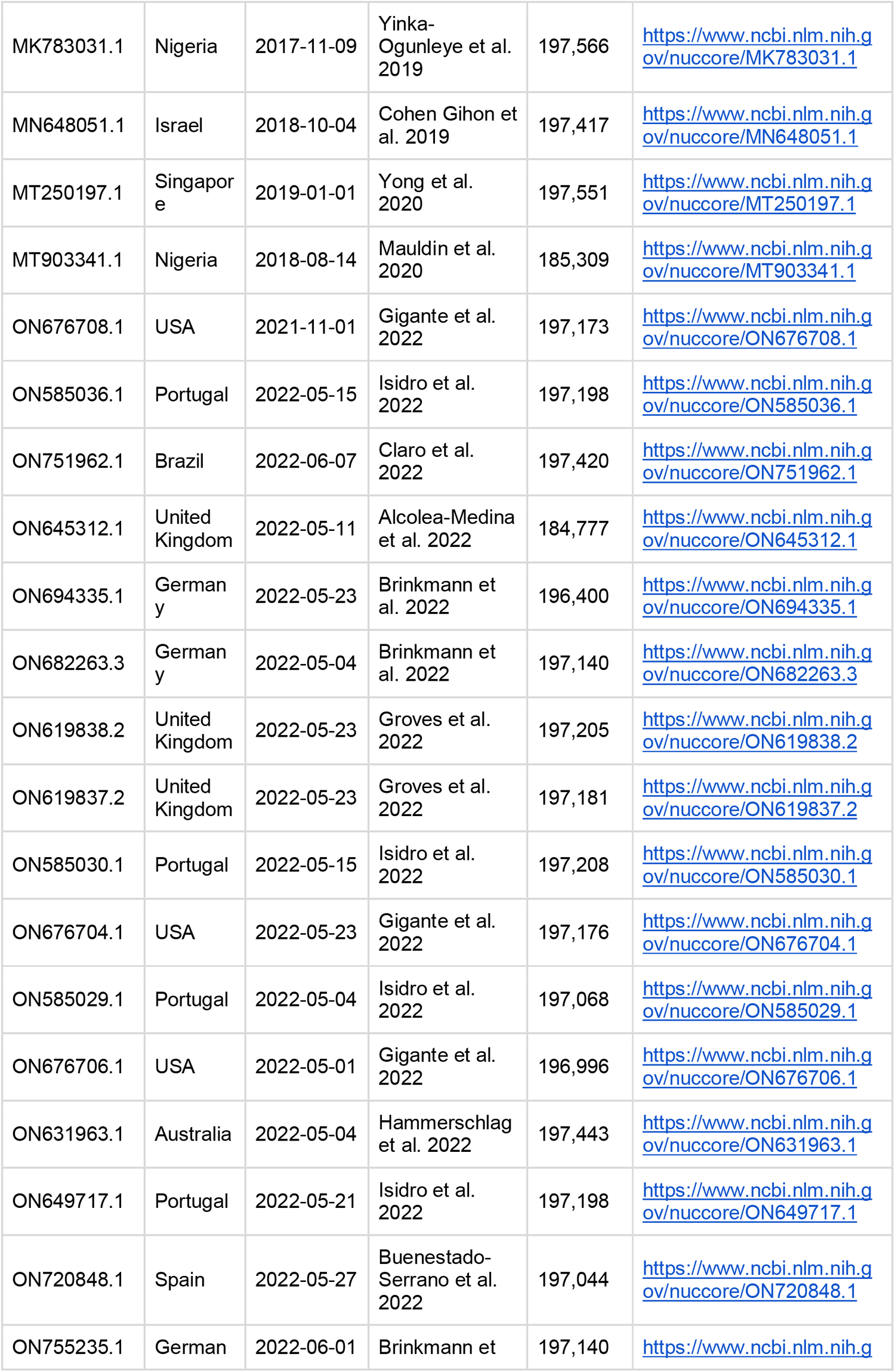
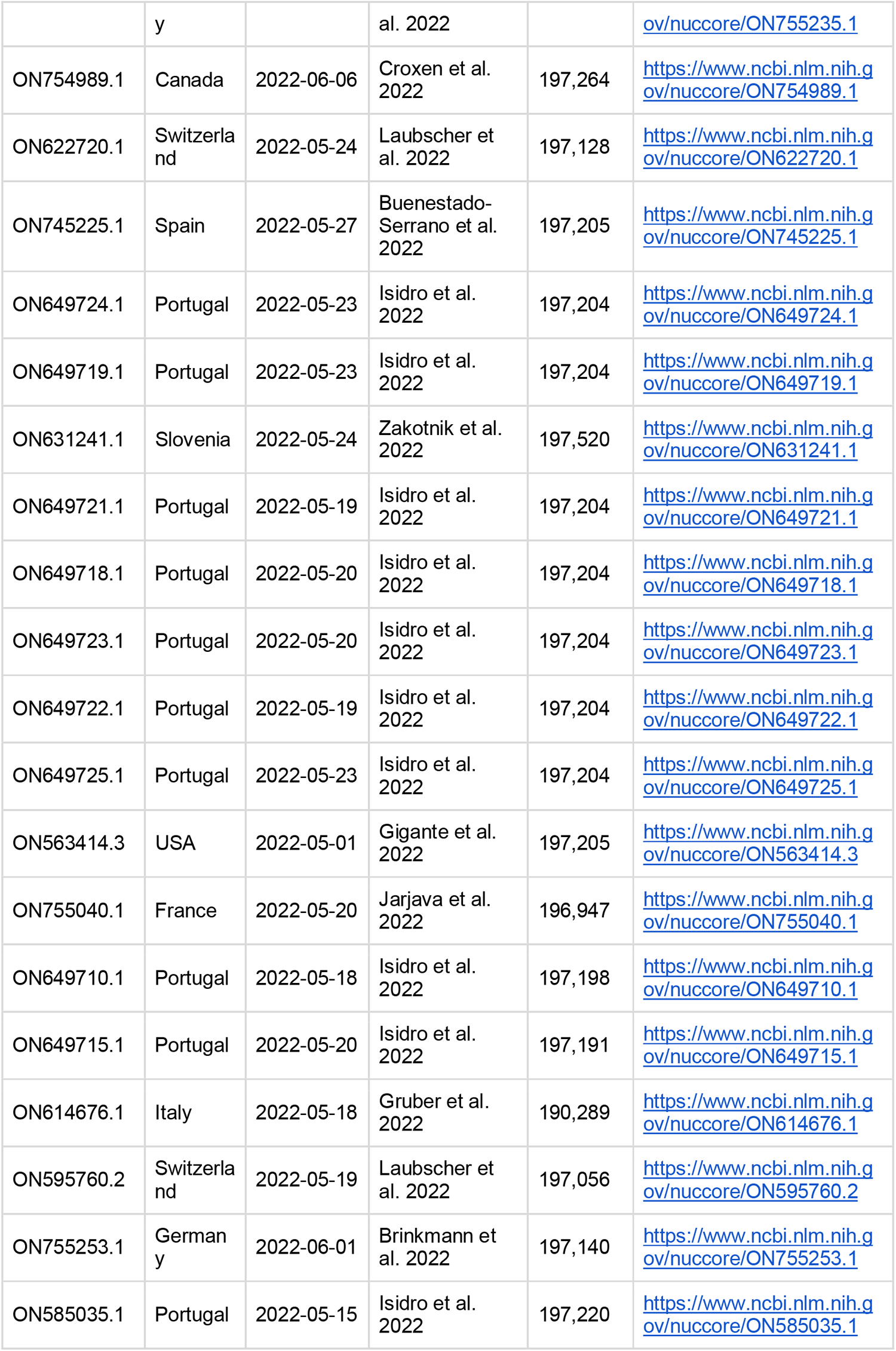
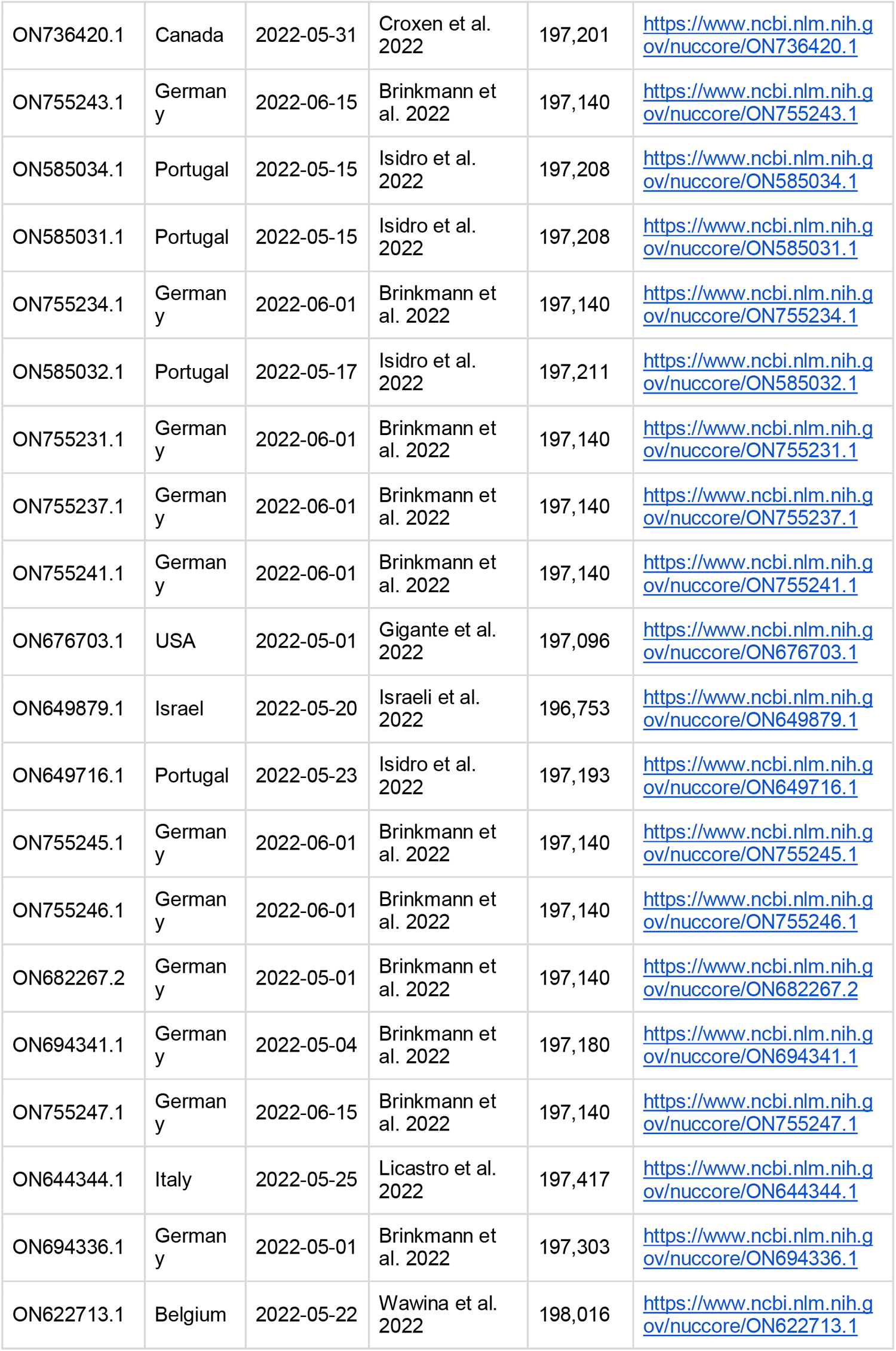
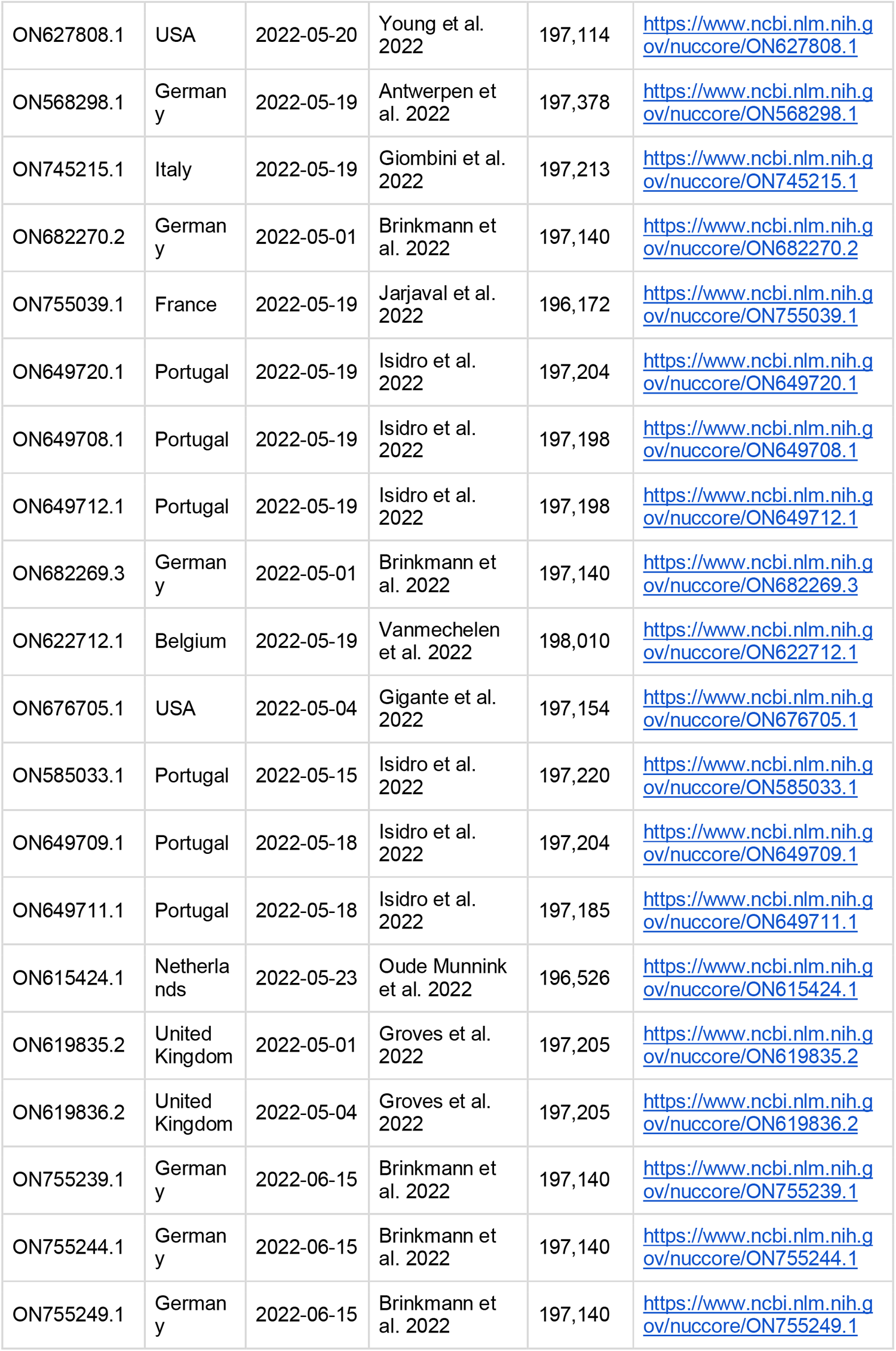
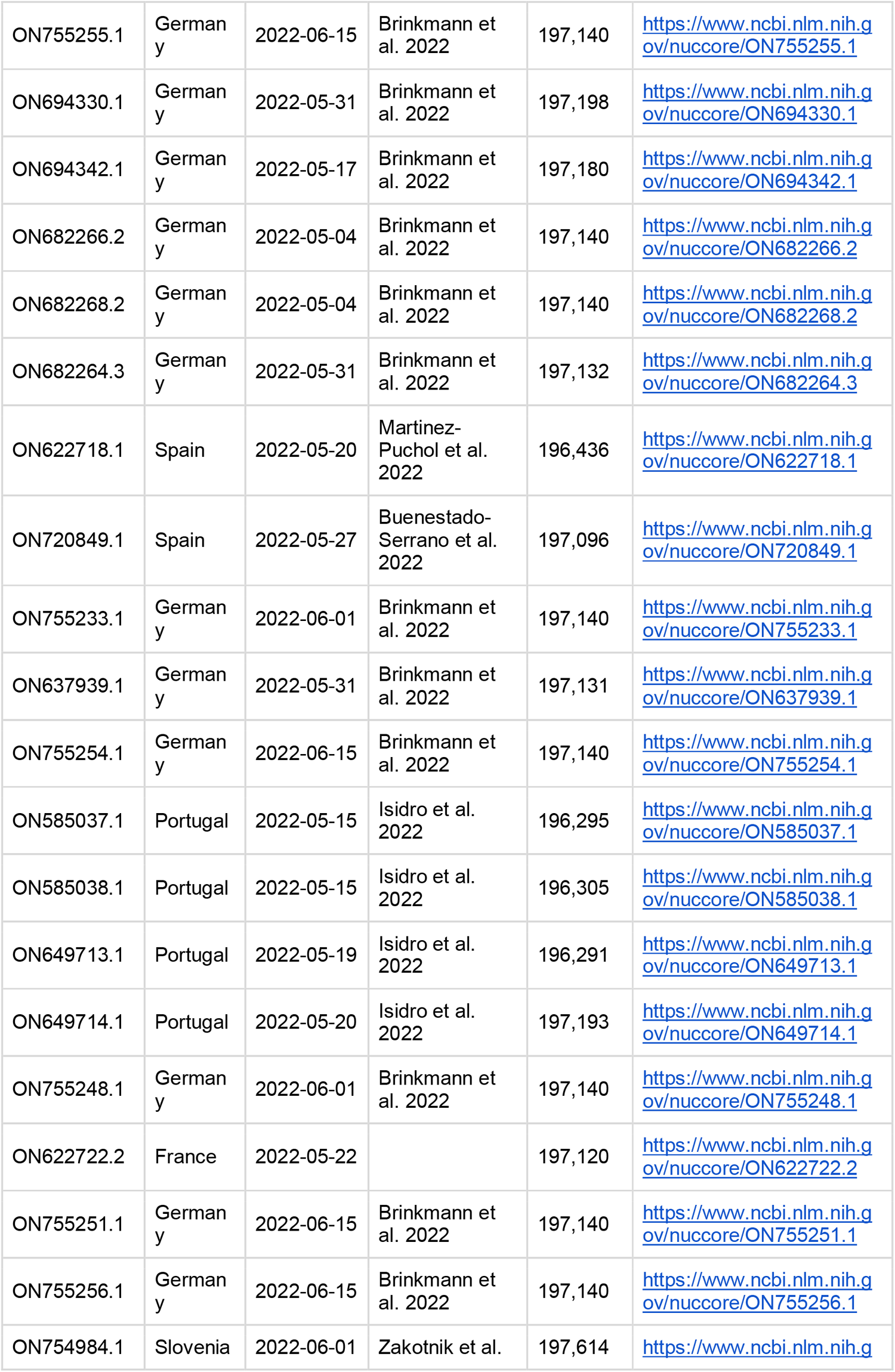
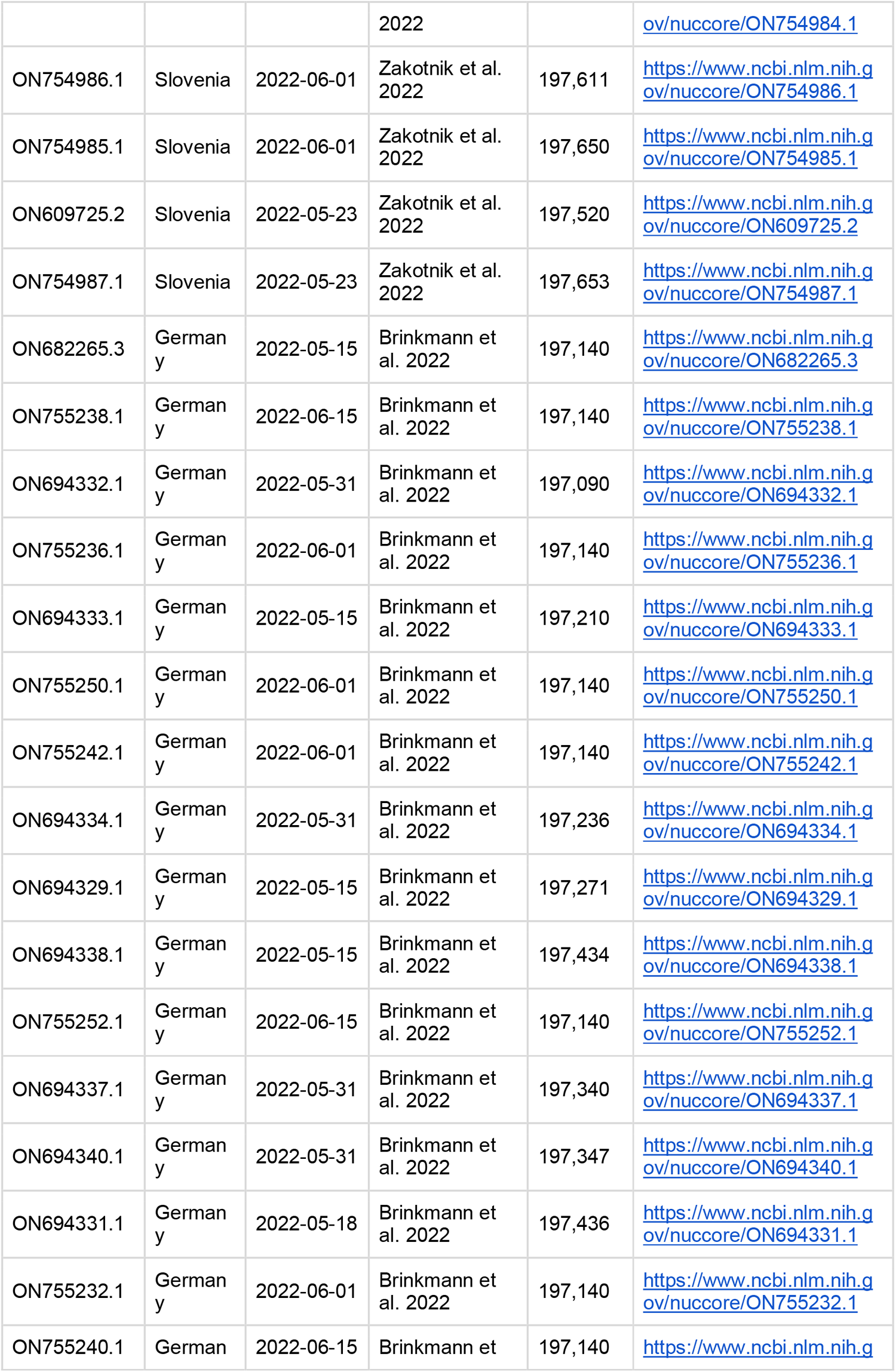
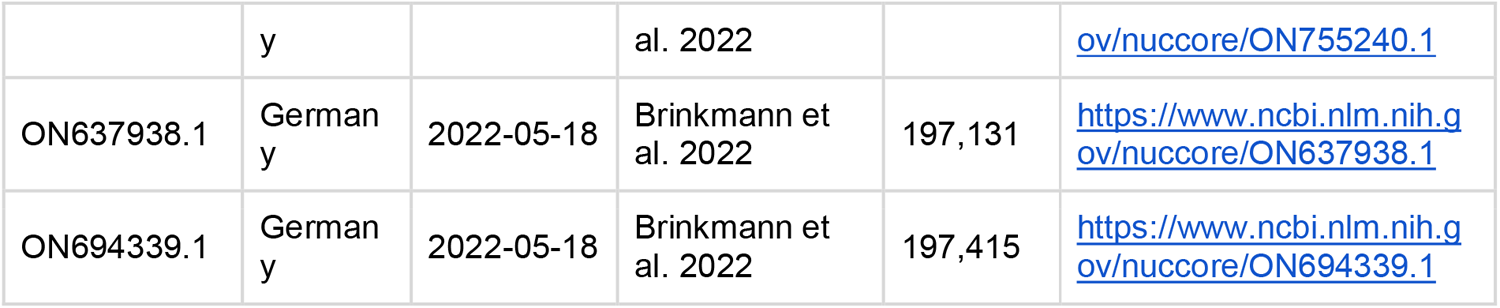
Acknowledgement and accession numbers used for phylogenetic analysis.

